# Accelerated MCDW-pCASL Using Subspace Low-Rank Reconstruction for Quantification of BBB Water Exchange and Permeability

**DOI:** 10.64898/2026.07.13.26357046

**Authors:** Zixuan Liu, Chenyang Zhao, Ziyang Huang, Fanhua Guo, Danny JJ Wang, Xingfeng Shao

## Abstract

**Purpose:** To develop an accelerated motion-compensated diffusion-weighted pseudo-continuous arterial spin labeling (MCDW-pCASL) method using a spatial subspace low-rank reconstruction method for efficient quantification of blood-brain barrier (BBB) water exchange (kw) and permeability (PSw).

**Methods:** An accelerated multidelay MCDW-pCASL sequence was developed to simultaneously encode intravascular and extravascular diffusion-weighted ASL signals across multiple post-labeling delays (PLDs). A spatial subspace low-rank reconstruction framework was optimized to enable joint estimation of cerebral blood flow (CBF) and BBB water exchange rate and permeability. Fourteen young healthy adults underwent test–retest scans (separated by ∼1 week) at 3T with both the accelerated MCDW-pCASL and a conventional diffusion-prepared (DP) pCASL sequence. Whole-brain, gray-matter, and white-matter CBF and kw values were quantified to assess test–retest repeatability and cross-method agreement. An additional cohort of 30 older adults underwent single-session MCDW and DP scans to evaluate age-related perfusion and BBB kw/PSw differences. Intraclass correlation coefficients (ICCs) were used to assess reliability and agreement.

**Results:** Accelerated MCDW-pCASL demonstrated excellent agreement with DP-pCASL for CBF (ICC = 0.89) and fair agreement for kw (ICC = 0.56). Test–retest repeatability of MCDW-pCASL was good for CBF, BBB kw and PSw (ICC ≈ 0.6). Across both sequences, younger subjects exhibited significantly higher CBF and kw compared with older adults.

**Conclusion:** Incorporating a spatial low-rank subspace reconstruction enables accelerated MCDW-pCASL acquisition with reliable simultaneous quantification of CBF, BBB kw and PSw. Clinical applications of this method for assessing perfusion and BBB function are warranted.

## 1. Introduction

The blood–brain barrier (BBB), as a highly selective semipermeable interface, is essential for maintaining central nervous system homeostasis, and its structural integrity relies on a sophisticated architecture of tight junctions between endothelial cells, pericytes, and astrocyte end-feet (1). Research indicates that the age-related degradation of these microscopic elements, such as the downregulation of tight junction proteins, would trigger subtle yet detrimental alterations in BBB permeability (2). This increased leakage facilitates the infiltration of systemic neurotoxic molecules into the brain parenchyma (3). It is also inextricably linked to neurovascular uncoupling and the progression of neurodegenerative diseases, including Alzheimer’s disease (AD) and related dementia (2), as well as motor neuron disease (MND)(4, 5). A thorough investigation into the integrity of the BBB and its evolutionary patterns during aging is of paramount importance for elucidating the underlying pathological mechanisms of such neurological disorders.

Dynamic contrast-enhanced (DCE) MRI is currently the most widely used approach to quantify BBB permeability to gadolinium-based contrast agents (GBCAs)(6). However, the moderate sizes of gadolinium chelates (∼0.55 kDa) and their predominantly paracellular pathway across the BBB limit the sensitivity of DCE MRI for detecting early or subtle BBB impairments (2). In addition, repeated use of GBCAs is associated with concerns regarding gadolinium deposition in the brain (7, 8), and their use is contraindicated or restricted in patients with impaired renal function (9). During recent years, several variants of arterial spin labeling (ASL) techniques have been introduced, including diffusion-weighted ASL (10) and multi-TE ASL (11), which indicate water as an endogenous contrast agent to assess BBB. Because of its small molecular size (∼18Da)(12) and multiple pathways for water to cross the BBB, including passive diffusion (13), co-transport by ion-pumps and aquaporin-4 (AQP4)–mediated transport (12, 14), these emerging methods can potentially detect early and/or subtle changes in BBB function and integrity during disease processes.

Diffusion prepared pseudo-continuous ASL (DP-pCASL) has been proposed to quantify the BBB water exchange rate (kw)(15), showing promising results of kw changes with aging (16), Alzheimer’s disease (17, 18) and cerebral small vessel disease (19). The diffusion preparation module with spoiling gradients to preserve the Carr-Purcell-Meiboom-Gill (CPMG) condition results in 50% signal loss (20). Consequently, relatively low spatial resolution (e.g. 3.5×3.5×8mm^3^) was applied in DP-pCASL to compensate for the SNR loss (15). Recently, a Motion Compensated Diffusion Weighted (MCDW) GRASE pCASL sequence has been proposed to improve the SNR and spatial resolution at multiple PLDs (21). By applying a two-compartment single-pass approximation (SPA) model with an additional venous compartment, MCDW-pCASL allows the simultaneous quantification of multiple hemodynamic and permeability parameters, including cerebral blood flow (CBF), kw, permeability-surface area product (PSw) and extraction fraction (Ew) of water. Compared with DP-pCASL, MCDW-pCASL offers an approximately three-fold increase in temporal signal-to-noise ratio (tSNR) allowing improved spatial resolution (∼iso-3.5mm). The kw measurements by MCDW-pCASL is highly consistent with those by DP-pCASL (ICC = 0.82), and the PSw measurements by MCDW-pCASL are comparable to literature values reported using the gold-standard of ^15^O-water PET (22). Despite these advantages, clinical adoption of MCDW-pCASL remains challenging due to its relatively long acquisition time (∼35 minutes) and the susceptibility of permeability measurements to noise-induced bias at the individual subject level. In addition, the performance and reliability of MCDW-pCASL in aging populations, where prolonged arterial transit delays are more prevalent, have yet to be established.

In this study, we propose a low-rank subspace denoising framework that enables accelerated MCDW-pCASL acquisitions by exploiting the intrinsic spatiotemporal redundancy of diffusion-weighted ASL signals. By projecting voxel-wise signals onto a compact subspace learned from combined simulated and in vivo data, the proposed approach improves the robustness of model estimation of BBB parameters against noise. The performance of the proposed method was evaluated using simulation, test–retest experiments, and comparison with standard DP-pCASL in both young and older participants.

## 2. Methods

### 2.1 Low-Rank Subspace Denoising

We propose a subspace low-rank imaging for water exchange rate (SLIWER) framework (23), which applies singular value decomposition (SVD) to capture the dominant temporal components of MCDW-pCASL signals. The method operates on label and control measurements acquired across 5 PLDs, with and without diffusion weighting, resulting in a total of 20 temporal samples per voxel (5 PLDs × label/control × without/with diffusion-weighted acquisitions). For each voxel, these measurements were arranged into a 20-point temporal signal vector representing the evolution of the MCDW-pCASL signal across PLDs and diffusion conditions.

To generate a representative signal subspace, in vivo MCDW-pCASL data were combined with simulated signals spanning a wide range of physiological parameters, including tissue T1 (0.5–1.6 s), CBF (1–120 ml/100 g/min), ATT ( 0.5–2.5 s), and kw (10–400 min⁻¹). Tissue T1 effects were explicitly incorporated into the simulations to account for signal modulation induced by background suppression pulses. Simulated signals were generated by sampling the physiological parameter ranges to cover diverse perfusion and permeability conditions. In addition, voxel-wise temporal signals from motion-corrected in vivo datasets were incorporated to capture realistic signal variations that may not be fully represented by the simulations.

Figure 1A illustrates the structure of the training database, which consists of simulated signals together with motion-corrected MCDW-pCASL data from five PLDs acquired in 38 subjects (24). All signals were organized into a signal matrix, where each column corresponds to a temporal signal vector and each row represents one time series of measurements across simulations and subjects. Singular values obtained from applying SVD to the combined simulated and in vivo datasets are shown in Figure 1B. A singular-value threshold of 0.2% of the maximum singular value was used to define the retained subspace, corresponding to 13 temporal components. This threshold was empirically determined based on the optimal balance between minimizing the limits of agreement of test-retest scans and preserving correlations among the tested denoising strategies and neighboring ranks, as shown in Supplementary Figures S1 and S2. Accordingly, the threshold >= 0.0035 was retained as the dominant signal subspace. The corresponding temporal basis functions are shown in Figure 1C, while the spatial coefficient maps derived from one representative subject are displayed in Figure 1D. The low-rank structure arises from the strong temporal correlations of ASL signals governed by perfusion kinetics and diffusion attenuation across PLDs, allowing the high-dimensional measurement space to be efficiently represented in a compact signal subspace.

**Figure 1.**
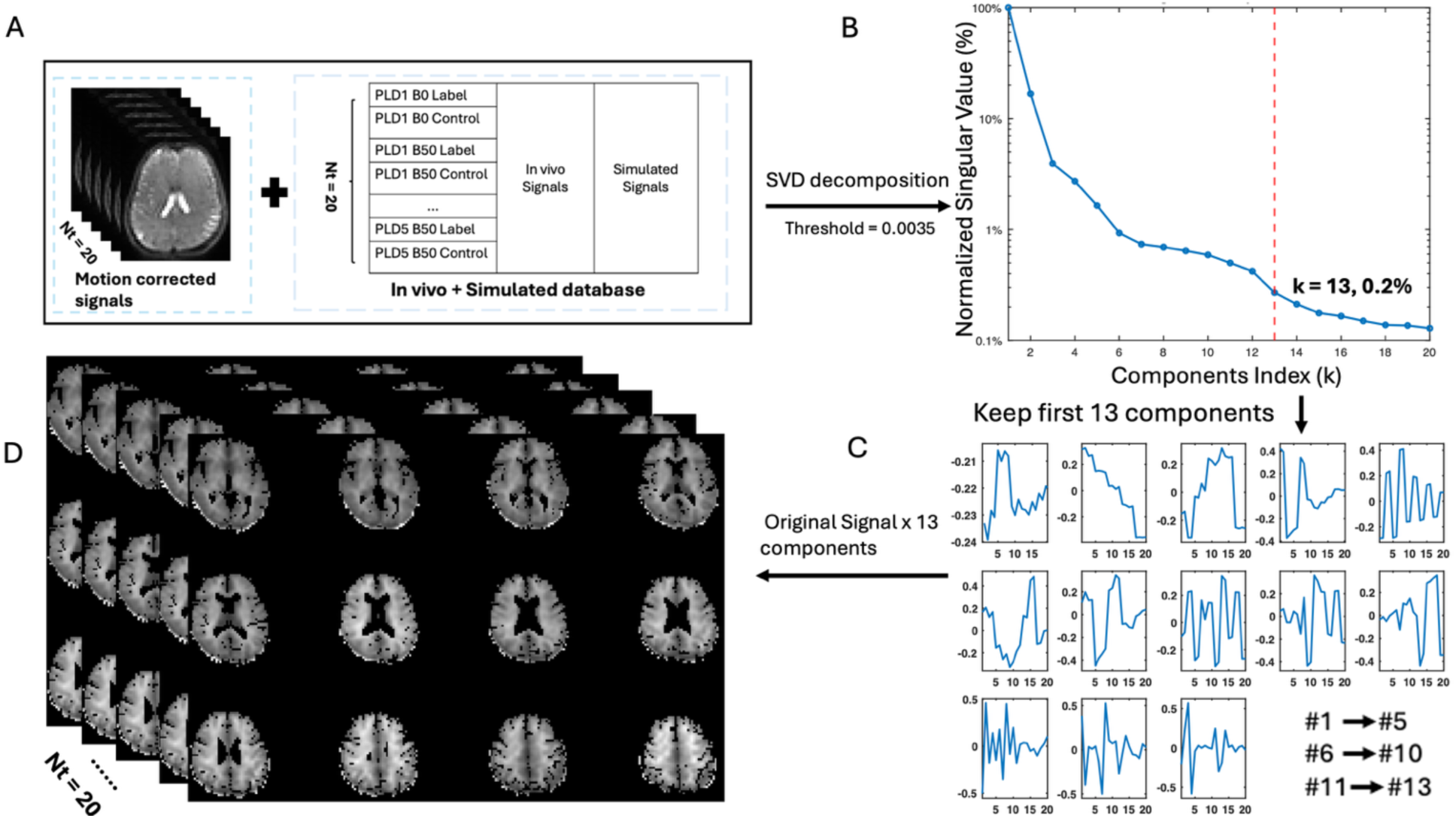
Subspace Low-Rank Imaging Method (SLIWER). (A) Displays the combination of motion-corrected subject data with the reference database. (B) Singular values obtained from the SVD of combined simulated and in-vivo MCDW-pCASL signals across 20 time points. Components with singular values smaller than 0.2% were considered as noise, and the first 13 components were selected as the major basis functions. (C) Extracted basis functions corresponding to the first 13 components. (D) Representative coefficient maps from one subject associated with these basis functions.

### 2.2 Monte Carlo Simulation

To demonstrate the denoising effect of SLIWER, a Monte Carlo simulation was applied to evaluate the robustness and accuracy of kw, as well as Ew and PSw estimations, from noisy MCDW-pCASL data. MCDW-pCASL signals without noise were first generated using a two-compartment SPA model with 5 PLDs (21), with a labeling duration of 1.87 s, yielding a total of 20 temporal samples per voxel, same as the in vivo imaging protocol. Physiological parameters were sampled over wide ranges, including brain tissue T1 (0.8–1.4 s), diffusion coefficient (0.7 – 1.0 x 10^-3^ mm^2^/s), CBF (20–100 ml/100 g/min), ATT (1.0–2.2 s), and kw varied from 60 to 160 min^-1^ in increments of 10 min^-1^. This range was chosen to reflect the intrinsic variability of physiological parameters, and the simulation was designed to assess whether the proposed method could robustly estimate a range of kw/Ew/PSw values rather than fitting a single fixed ground-truth value.

To simulate realistic in vivo conditions, structured signal complexity was introduced before noise addition, including subject-level temporal modulation to mimic inter-subject variability. Specifically, the simulated signals were divided into 10 virtual subjects, each assigned a distinct smooth temporal modulation pattern generated by Gaussian filtering across time points and a random amplitude scaling factor between 0.85 and 1.25, PLD-dependent labeling efficiency variations with a standard deviation of 15% across PLDs, low-frequency temporal drift modeled as a sinusoidal modulation with a maximum amplitude of 10%, spatially correlated low-rank fluctuations constructed from eight spatiotemporal components scaled to 25% of the global signal standard deviation, voxel-wise amplitude scaling with a standard deviation of 25%, and sporadic motion-related perturbations introduced at four randomly selected time points with amplitudes of 30% of the signal standard deviation and attenuated residual effects in adjacent frames. Multiple noise sources were then added to approximate both unstructured measurement noise and structured temporal–spatial fluctuations commonly observed in ASL acquisitions. These included zero-mean Gaussian noise with a standard deviation equal to 40% of the global signal standard deviation, low-rank temporal–spatial noise generated from 16 components and scaled to 15% of the global signal standard deviation, Rician magnitude noise with a noise scale equal to 10% of the global signal standard deviation, and additional spatially correlated noise generated from 20 components and scaled to 8% of the global signal standard deviation.

A total of 1000 Monte Carlo repetitions were performed, each with independently generated realizations of signal complexity and noise. For each Monte Carlo realization, kw was estimated from both noisy and denoised datasets using the same linear regression of logarithm (LRL) fitting procedure applied to the in vivo data. The estimation was performed voxel-wise using the simulated MCDW-pCASL signals across all PLDs and diffusion conditions. Based on the estimated kw and the reconstructed ASL difference signals, Ew was subsequently calculated using the ratio of ASL signals at the two latest PLDs with and without diffusion-weighting, and PSw was derived from Ew and the correspondingly estimated CBF according to the estimation model described below. For the voxel–wise Bland-Altman analysis, parameters estimated were first averaged across 1000 Monte Carlo repetitions for each voxel. The voxel-wise averaged estimates were then compared with the corresponding noise-free reference values, with the mean of estimated and reference values on the x-axis and their difference on the y-axis.

### 2.3 MRI Experiments

A total of 14 healthy young subjects (here referred as the young group) participated in this study (5F/9M, age=28.3±9.4 years) who underwent MRI scans on a 3T Prisma system (Siemens Healthineers, Erlangen, Germany) using a 32-channel head coil. All subjects underwent test-retest scans within two weeks that included both MCDW-pCASL and DP-pCASL scans. In addition, 30 elderly subjects (16F/14M, age= 69.83 ± 5.34 years) underwent MCDW-pCASL scans on the same 3T Prisma system. All subjects provided written informed consent according to a protocol approved by the Institutional Review Board of the University of Southern California.

Imaging parameters for MCDW-pCASL were: FOV=224 mm, matrix size=64×64, 36 slices with 20% oversampling, isotropic resolution of 3.5×3.5×3.5 mm³, EPI factor=63, turbo factor=15, and 3-fold acceleration along the partition direction (25). The bandwidth was 2232 Hz/pixel, and TE was 47.7 msec. Label and control duration was 1870 msec. Background suppression was achieved using a pre-saturation pulse followed by two hyperbolic secant (HS) inversion pulses (26). Six label/control pairs were acquired at b=0 or 40.4 sec/mm² across five PLDs (1500, 1800, 2100, 2400, and 2700 msec), corresponding to TRs of 4300, 4600, 4900, 5200, and 5500 msec, respectively. Acquisition times for each PLD with six measurements ranged from 2 min 03 sec to 2 min 38 sec. A separate M0 scan was acquired in 15 sec, resulting in a total scan time of approximately 12 min.

For comparison, a DP-pCASL scan was performed using matched in-plane resolution and FOV (12 slices without acceleration, thickness = 8 mm)(15). The DP-pCASL protocol employed background suppression and a separate M0 acquisition, and a two-stage SPA modeling framework with TGV regularization was used to derive ATT, CBF, and kw within a total scan time of approximately 10 min. ATT was estimated using FEAST acquisitions (27) at PLD = 900 ms with 15 repetitions for each of the two diffusion weightings (b = 0 and 10 sec/mm²), kw was quantified at a longer PLD of 1800 ms using diffusion-weighted (b = 50 sec/mm²) and non-diffusion-weighted (b = 0 sec/mm²) signals with 20 repetitions per condition; while CBF was calculated from the M0 image and the corresponding non-diffusion-weighted (b = 0) ΔM acquired at PLD = 1800 ms.

### 2.4. Image Processing and Data Analysis

#### 2.4.1 Pre-processing

All image processing and data analysis were conducted on a stand-alone workstation using in-house scripts and algorithms implemented in MATLAB (The MathWorks, Inc., Natick, MA). Accelerated MCDW-pCASL data were reconstructed using the Berkeley Advanced Reconstruction Toolbox (28). Rigid-body motion correction was applied to both control and label images, which were subsequently co-registered to the corresponding M0 images using SPM12 (Wellcome Trust Centre for Neuroimaging, UCL, London, UK). Perfusion-weighted images were generated by pairwise subtraction of control and label images, and residual temporal fluctuations arising from motion and physiological noise were reduced using a principal component analysis (PCA)-based approach (29).

Gray matter (GM) and white matter (WM) masks derived from T1-weighted image segmentation were co-registered to the M0 images using SPM12 and used for tissue-specific analysis. SLIWER denoising was applied using singular value decomposition (SVD) to construct a low-rank temporal subspace for MCDW-pCASL signals. The SLIWER database was generated by combining in-vivo label/control repetitions from 38 subjects with the Monte Carlo simulated signal evolutions described above (16). The simulated signals accounted for approximately 10% of the database and were included to increase physiological variability in the training subspace. SVD was then performed on the combined database to extract the dominant temporal components of the label/control signal evolution. For each voxel, the measured MCDW-pCASL signal was projected onto the retained SVD basis and reconstructed for denoising.

#### 2.4.2 kw quantification

The denoised signals were then used for voxel-wise estimation of kw, Ew, and PSw. CBF and ATT were estimated using standard perfusion model fitting of 5-PLD data (30). BBB kw was estimated using a voxel-wise Linear Regression of Logarithm Signal (LRL) based on the decay of capillary-weighted perfusion signals across PLDs (21). Under the two-compartment SPA model, the capillary-weighted perfusion signal *M_c_*(*t*) can be expressed as:

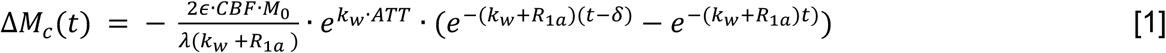

where *ε* is the labeling efficiency. By taking the logarithm of both sides, the exponential decay term can be linearized as:

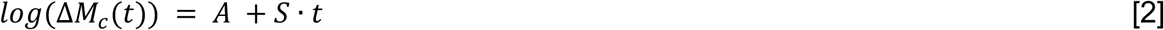

where A is the intercept, and S is the slope of the linear regression. This allows kw to be efficiently estimated on a voxel-wise basis directly from the slope of the regression line:

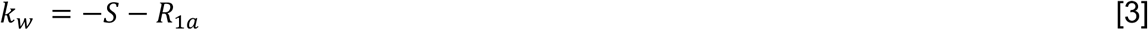

To enhance fitting robustness, all possible PLD subsets containing at least three time points were evaluated. Based on the voxel-wise ATT, PLD subsets were dynamically adjusted: for voxels with ATT longer than 1.9 s, the two early PLDs (1.5 and 1.8sec) were excluded since the labeled blood hasn’t reached capillaries, whereas for voxels with ATT longer than 1.6s but shorter than 1.9 s, the first and last PLDs were excluded; otherwise, the fifth PLD was preferentially excluded because its lower temporal SNR could introduce greater uncertainty into kw estimation (shown in Figure S3). This design accounts for the low tSNR at the longest PLD, which may otherwise introduce instability in slope estimation. Multiple fitting strategies were applied for comparison, including voxel-only fitting and mixed voxel–neighborhood fitting with varying weighting ratios. Candidate fit algorithms were evaluated based on predefined physiological and statistical criteria. Fits with excessive residual error (RMSE ≥ 1.0 in the log-signal domain) were excluded. To improve robustness against outliers, median absolute deviation (MAD)-based filtering was applied, whereby signal points deviating more than three times the MAD from the median were replaced by the median prior to regression. If no valid voxel-wise solution was identified, kw was estimated using the global prior. tSNR measurements for each PLD with and without diffusion weighting are summarized in Figure S3.

#### 2.4.3 Ew and PSw quantification

The water exchange fraction Ew was estimated from perfusion signals acquired at the 2 latest PLDs based on the assumption that the tissue fraction can approximate water extraction when the PLD is sufficiently long to allow labeled blood arrival and capillary exchange, approximately when PLD > ATT + *τ_c_*. Following the previous MCDW-pCASL framework, whole-brain ATT was estimated to be 1250.0 ms in GM, and 1311.3 ms in WM, and the reported *τ_c_* value was approximately 1400 ms (21). Under this assumption, the two longest PLDs in the present study, 2400 and 2700 ms, were used for Ew estimation, as our prior work showed that tissue fractions measured at comparable late PLDs differed from the model-derived Ew by only approximately 1–3% (21). Specifically, *Ew* on each voxel was calculated as:

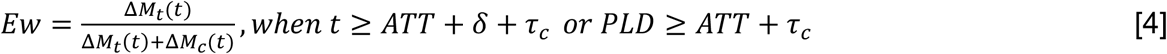

where Δ*M_t_* and Δ*M_c_* indicate the tissue and capillary components of the ASL signal, respectively, averaged across the last two PLDs. Then, the permeability–surface area product (*PS_w_*) was derived as:

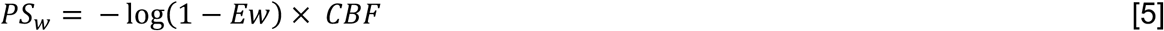

To improve consistency with the model assumption mentioned above, only voxels with estimated ATT below 1600 ms were included in the calculation of Ew and PSw.

#### 2.4.4 DP-pCASL processing

A DP-pCASL toolbox developed using custom MATLAB scripts by the LOFT lab at USC was used to analyze all DP-pCASL data. CBF and kw maps obtained from DP-pCASL and MCDW-pCASL were compared and the agreement between whole-brain–averaged CBF and kw values across subjects measured by the two techniques was quantified using the intraclass correlation coefficient (ICC).

#### 2.4.5 Statistical analysis of age effect

Age effects on whole-brain kw and CBF were evaluated using independent two-sample t-tests between young and elderly groups for DP-pCASL and MCDW-pCASL, respectively. For MCDW-derived BBB permeability metrics (Ew and PSw), age-related differences were similarly assessed using independent two-sample t-tests between young and elderly subjects. Normality of the data distribution was verified before parametric testing. Statistical significance was defined as two-tailed p < 0.05.

## 3. RESULTS

### 3.1 Simulation-Based Evaluation of Low-Rank Subspace Denoising

Figure 2 illustrates the performance of the SVD-based denoising method in estimating parameters from Monte Carlo simulated noisy ASL data. The simulation results demonstrate a clear improvement in the estimated kw distribution, with greater improvements of Ew and PSw distributions toward the ground truth following denoising. Specifically, the noisy dataset yielded an estimated mean kw of 129.8 ± 40.2 min^-1^, with a positive bias (2.89) relative to the ground-truth mean of 121.5 ± 28.5 min^-1^. However, there are large variations in the estimated kw using noisy data relative to the ground-truth, manifested by a normalized RMSE (NRMSE) of 0.29 and a wide BA width of 135.69 min^-1^. In contrast, the denoised dataset yielded an estimated mean kw of 130.0 ± 27.7 min^-1^ with a comparable positive bias of 3.07 but significantly reduced NRMSE of 0.22 and a narrower BA width of 104.62 min^-1^ (Figure 2A&B). For Ew, the reference value was 85.07% ± 16.61%, while the denoised and noisy estimates were 77.47% ±7.6 % and 71.14% ± 11.5 %, respectively. The BA bias was -7.6% and -12.4%, the NRMSE was 0.41 and 0.65, and the BA width was 36.6% and 50.14% for the denoised and noisy data, respectively (Figure 2C&D). For PSw, the reference value was 130.73± 69.65 ml/100 g/min, while the denoised and noisy estimates were 134.75 ± 69.2 and 148.96 ± 88.5 ml/100 g/min, respectively. The corresponding BA bias was 4.02 and 18.24 ml/100 g/min, the NRMSE was 0.12 and 0.28, and the BA width was 159.94 and 361.32 for the denoised and noisy data, respectively. The accuracy of parameter estimation from the Monte Carlo simulations is further summarized in Table 1.

**Figure 2.**
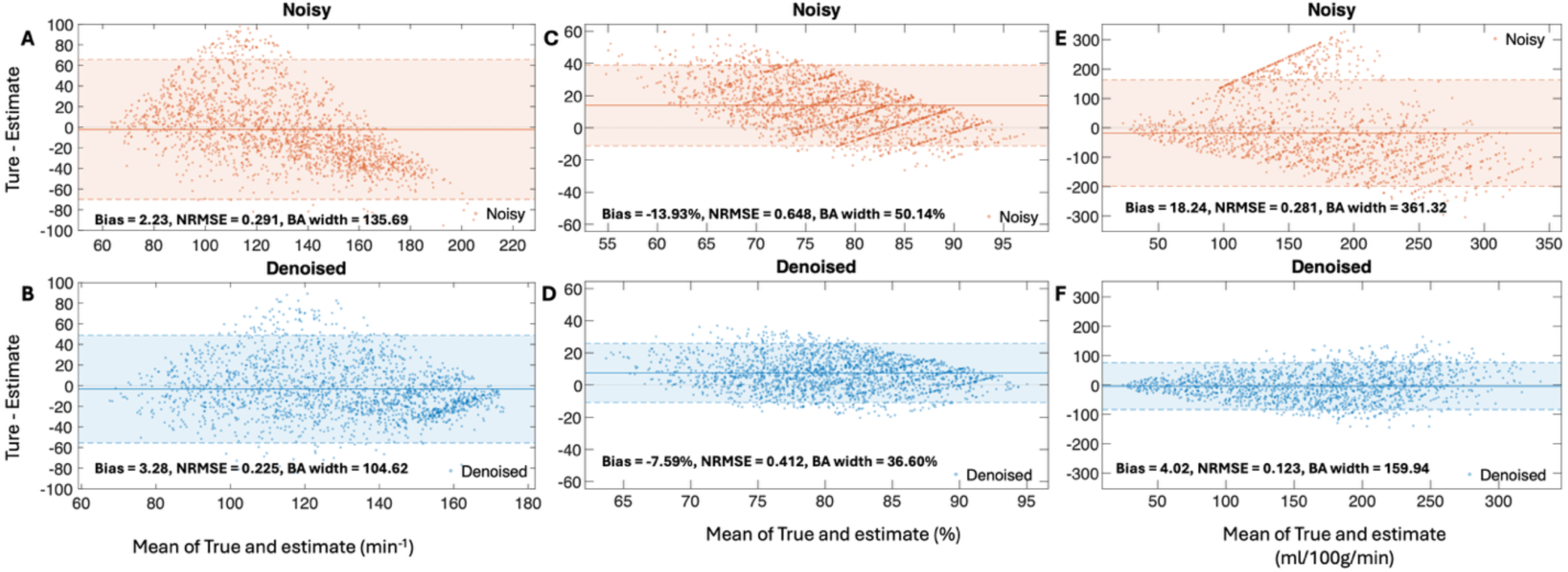
Bland–Altman analysis of kw, Ew, and PSw estimates from noisy and denoised datasets. Panels A–B show kw, panels C–D show Ew, and panels E–F show PSw. The upper row shows the noisy results, and the lower row shows the denoised results. The y-axis represents the difference between the true and estimated values, and the x-axis represents the mean of the true and estimated values. Solid horizontal lines indicate the mean bias, and dashed lines indicate the 95% limits of agreement. Panels A and B show kw with Bias = 2.23, NRMSE = 0.29, BA width = 135.69 for noisy data and Bias = 3.28, NRMSE = 0.22, BA width = 104.62 for denoised data. Panels C and D show Ew with Bias = −13.93%, NRMSE = 0.65, BA width = 50.14% for noisy data and Bias = −7.59%, NRMSE = 0.41, BA width = 36.60% for denoised data. Panels E and F show PSw with Bias = 18.24, NRMSE = 0.28, BA width = 361.32 for noisy data and Bias = 4.02, NRMSE = 0.12, BA width = 159.94 for denoised data.

**Table 1.** Parameter estimation results for kw, Ew, and PSw obtained from denoised and noisy datasets across 1000 Monte Carlo repetitions. Estimation accuracy is quantified using normalized root mean square error (NRMSE) and BA bias from noise-free simulations. Improvement indicates the percentage reduction in MAE or RMSE after SLIWER denoising.

| Parameter | Metric | Clean | Denoised | Noisy |
| --- | --- | --- | --- | --- |
| kw | Mean ± SD (min<sup>-1</sup>) | 121.5 ± 28.5 | 130.0 ± 27.7 | 129.8 ± 40.2 |
|  | NRMSE | — | **0.22** | 0.29 |
|  | BA Bias | — | 3.07 | 2.89 |
| Ew | Mean ± SD (%) | 85.07% ± 16.61% | 77.47% ± 7.6% | 71.14% ± 11.5% |
|  | NRMSE | — | **0.41** | 0.65 |
|  | BA Bias | — | -7.6% | -12.4% |
| PSw | Mean ± SD (ml/100g/min) | 130.73 ± 69.65 | 134.75 ± 69.2 | 148.96 ± 88.5 |
|  | NRMSE | — | **0.12** | 0.28 |
|  | BA Bias | — | 4.02 | 18.24 |

### 3.2 Test-retest repeatability

Figure 3 shows the detailed test–retest results from 14 young subjects. Both kw and CBF showed good test–retest reproducibility using the MCDW sequence, with ICC values of 0.73 and 0.75 and CoV (coefficient of variation) values of 23.15% and 15.62%, respectively. For the permeability-related metrics, Ew showed a lower ICC of 0.35 and CoV of 4.15%, suggesting low inter-subject variability in Ew estimations. PSw showed fair repeatability, with an ICC of 0.60 and CoV of 16.0%.

**Figure 3.**
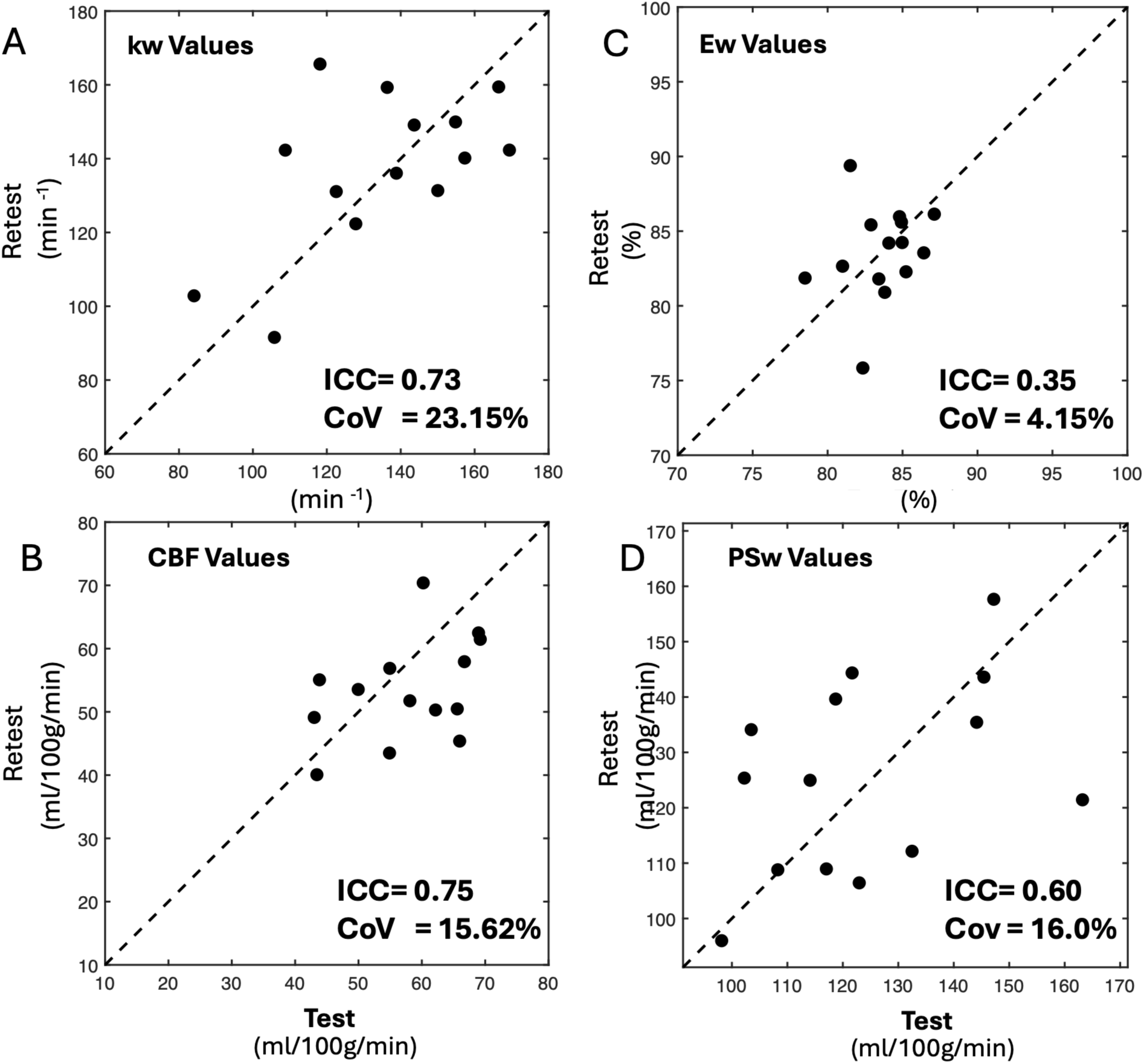
Scatter plots of test–retest whole-brain kw, CBF, Ew, PSw values across the 14 young subjects from MCDW scans, where the x-axis represents data from visit 1 and the y-axis represents data from visit 2. The ICCs were 0.73 for kw,0.75 for CBF, 0.35 for Ew, and 0.60 for PSw, respectively.

The quantitative results are summarized in Table 2, which reports the mean ± SD values and ICCs of CBF, kw, Ew, and PSw from MCDW-pCASL, as well as CBF and kw from DP-pCASL, across the whole brain, gray matter (GM), and white matter (WM).

**Table 2.**
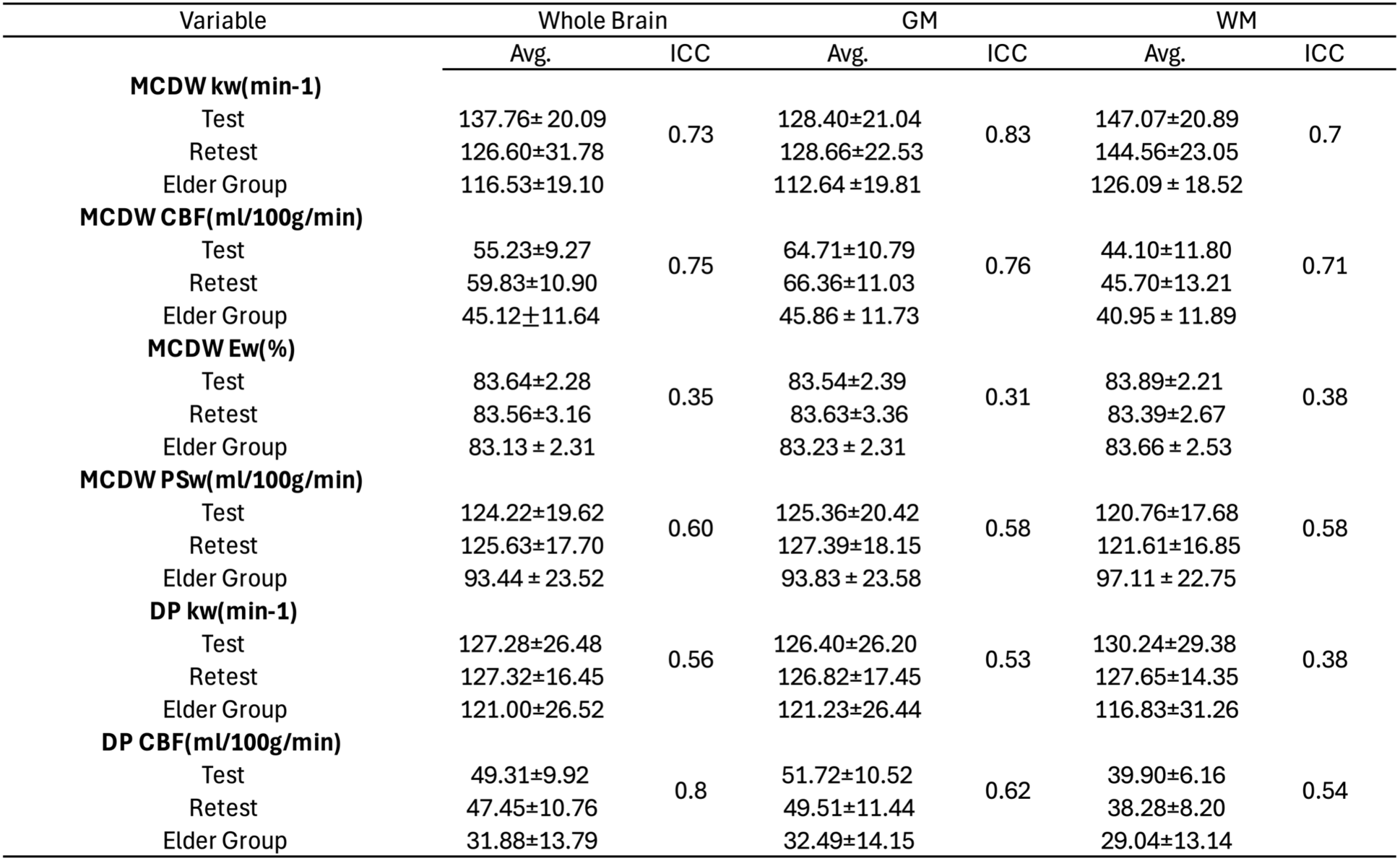
Mean values and ICCs of kw and CBF, Ew, and PSw in the whole brain, GM, and WM were measured using MCDW-pCASL and DP-pCASL from test–retest scans of 14 young and 30 older subjects (kw unit: min⁻¹; CBF unit: ml/100 g/min; Ew unit: %; PSw unit: ml/100 g/min ).

| Variable | Whole Brain |  | GM |  | WM |  |
| --- | --- | --- | --- | --- | --- | --- |
|  | Avg. | ICC | Avg. | ICC | Avg. | ICC |
| **MCDW kw(min-1)** |  |  |  |  |  |  |
| Test | 137.76±20.09 | 0.73 | 128.40±21.04 | 0.83 | 147.07±20.89 | 0.7 |
| Retest | 126.60±31.78 |  | 128.66±22.53 |  | 144.56±23.05 |  |
| Elder Group | 116.53±19.10 |  | 112.64±19.81 |  | 126.09±18.52 |  |
| **MCDW CBF(ml/100g/min)** |  |  |  |  |  |  |
| Test | 55.23±9.27 | 0.75 | 64.71±10.79 | 0.76 | 44.10±11.80 | 0.71 |
| Retest | 59.83±10.90 |  | 66.36±11.03 |  | 45.70±13.21 |  |
| Elder Group | 45.12±11.64 |  | 45.86±11.73 |  | 40.95±11.89 |  |
| **MCDW Ew(%)** |  |  |  |  |  |  |
| Test | 83.64±2.28 | 0.35 | 83.54±2.39 | 0.31 | 83.89±2.21 | 0.38 |
| Retest | 83.56±3.16 |  | 83.63±3.36 |  | 83.39±2.67 |  |
| Elder Group | 83.13±2.31 |  | 83.23±2.31 |  | 83.66±2.53 |  |
| **MCDW PSw(ml/100g/min)** |  |  |  |  |  |  |
| Test | 124.22±19.62 | 0.60 | 125.36±20.42 | 0.58 | 120.76±17.68 | 0.58 |
| Retest | 125.63±17.70 |  | 127.39±18.15 |  | 121.61±16.85 |  |
| Elder Group | 93.44±23.52 |  | 93.83±23.58 |  | 97.11±22.75 |  |
| **DP kw(min-1)** |  |  |  |  |  |  |
| Test | 127.28±26.48 | 0.56 | 126.40±26.20 | 0.53 | 130.24±29.38 | 0.38 |
| Retest | 127.32±16.45 |  | 126.82±17.45 |  | 127.65±14.35 |  |
| Elder Group | 121.00±26.52 |  | 121.23±26.44 |  | 116.83±31.26 |  |
| **DP CBF(ml/100g/min)** |  |  |  |  |  |  |
| Test | 49.31±9.92 | 0.8 | 51.72±10.52 | 0.62 | 39.90±6.16 | 0.54 |
| Retest | 47.45±10.76 |  | 49.51±11.44 |  | 38.28±8.20 |  |
| Elder Group | 31.88±13.79 |  | 32.49±14.15 |  | 29.04±13.14 |  |

Overall, the MCDW sequence demonstrated test–retest reliability comparable to that of the DP acquisition in the whole brain and GM/WM regions. At the whole-brain level, kw from MCDW showed consistent test/retest measurements (137.76 ± 20.09 / 126.06 ± 31.78 min⁻¹); which is higher than the repeatability of DP-pCASL (ICC = 0.56). MCDW-CBF likewise achieved comparable reliability (ICC = 0.75) compared to DP-CBF (ICC = 0.80), with closely matched test/retest results (55.23±9.27/59.83±10.90 ml/100g/min). Regionally, GM generally showed higher repeatability than WM. For MCDW-derived kw, the ICC was 0.83 in GM and 0.70 in WM. A similar pattern was observed for MCDW-derived CBF, with ICC values of 0.76 in GM and 0.71 in WM. Ew showed relatively low ICC values across regions, with ICCs of 0.35, 0.31, and 0.38 in the whole brain, GM, and WM, respectively, likely due to the small inter-subject variability. PSw showed fair repeatability, with ICC values of 0.60 in the whole brain and 0.58 in both GM and WM.

### 3.3 Comparison of MCDW vs. DP-pCASL

Figure 4 demonstrates the correlations of CBF and kw between MCDW and DP-pCASL, with orange squares representing the older group and blue triangles representing the young group. The kw values show fair agreement between DP and MCDW (ICC = 0.56) in whole brain, whereas CBF demonstrates an excellent agreement (ICC = 0.89). The correlations of CBF and kw in GM and WM are provided in Supplementary Figure S4, which shows patterns consistent with the whole-brain results.

**Figure 4.**
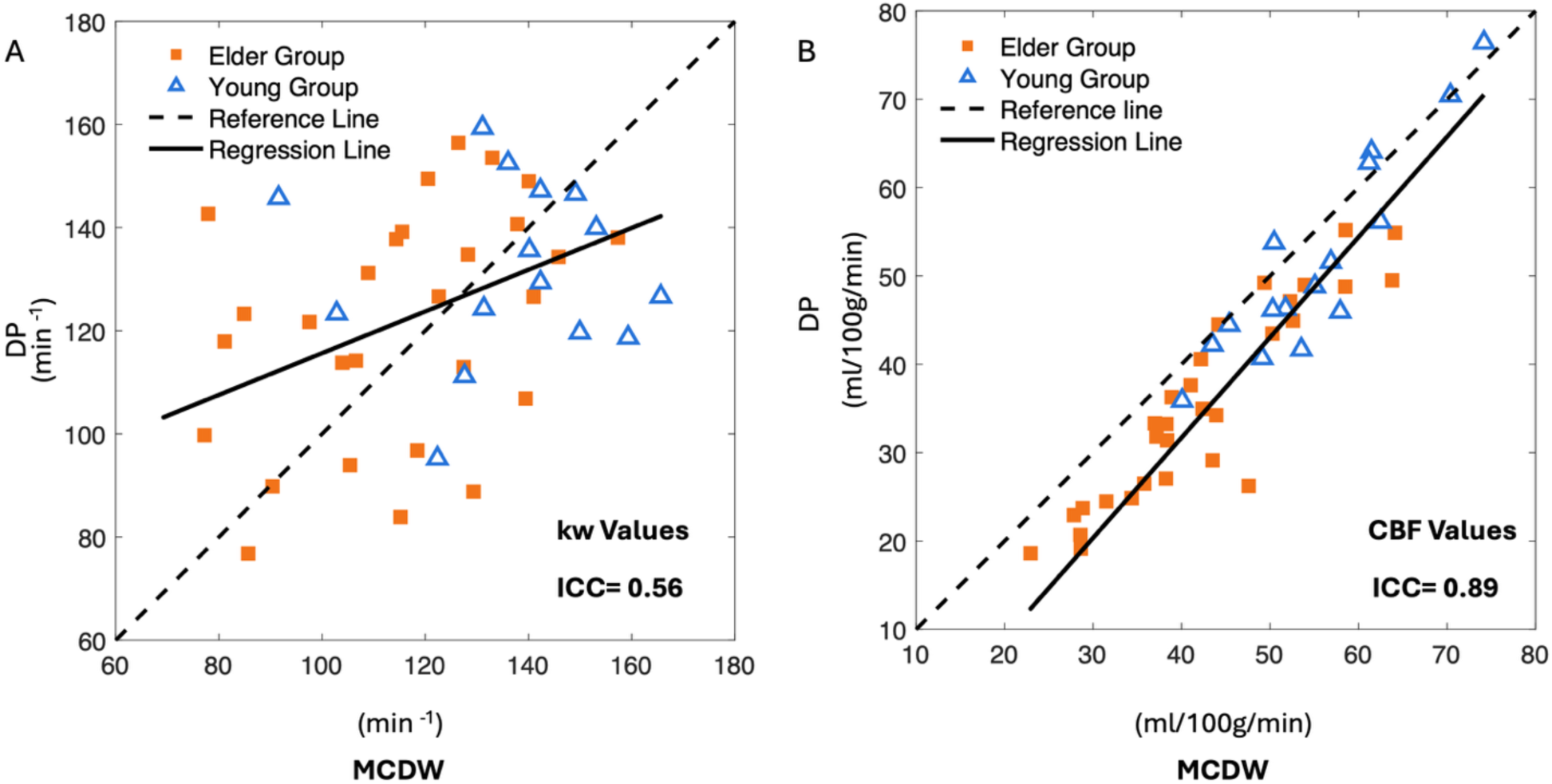
(A) and (B) present scatter plots showing correlations of whole-brain kw and CBF values across 14 young subjects (blue triangles) and 30 older subjects (orange squares) obtained from DP and MCDW scans, with correlation coefficients of ICC = 0.56 for kw, ICC = 0.89 for CBF.

Figure 5 displays representative slices of kw and CBF maps from a female subject aged from 25 to 30 years for both test and retest scans acquired using MCDW and DP-pCASL sequences, along with PSw and Ew maps obtained from the MCDW-pCASL sequence. Test results are shown at the top, and retest results at the bottom. All methods demonstrate relatively robust and consistent CBF and kw/Ew/PSw measurements across sessions.

**Figure 5.**
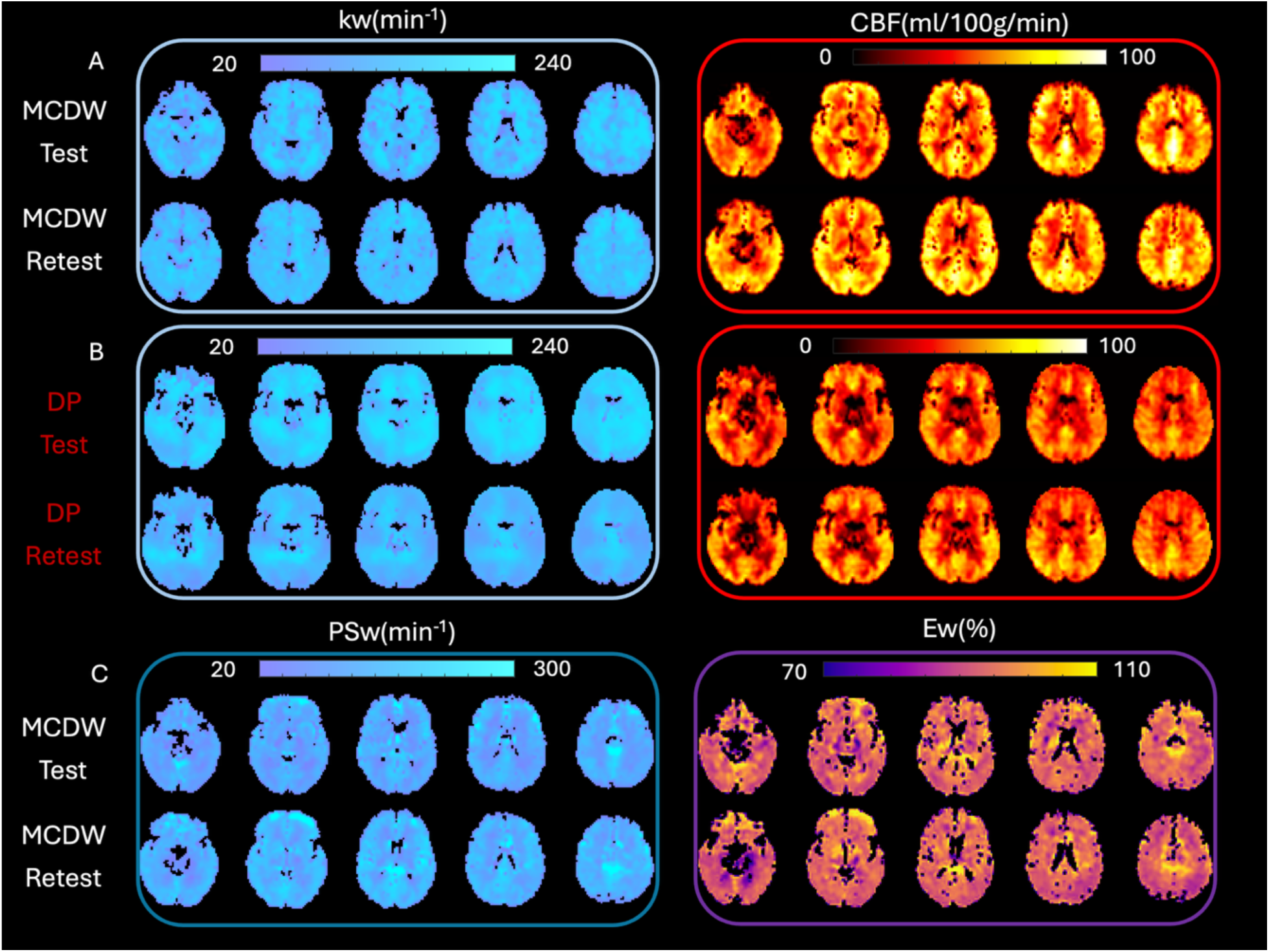
CBF, kw, and ATT maps show the test–retest results from a single subject (25-30 years old, female). (A–B) Whole-brain kw (blue squares) and CBF (red squares) maps from (A) MCDW-pCASL and (B) DP-pCASL, with test results displayed at the top and retest results at the bottom. (C) Whole-brain PSw (dark blue squares) maps and (D) Ew (purple squares) maps from MCDW-pCASL.

### 3.4 Evaluation of Age-Dependent Variations in kw, CBF, Ew, and PSw

Figure 6 displays the comparisons of CBF and kw/Ew/PSw measurements derived from MCDW-pCASL and DP-pCASL in elder and young subjects. As shown in Figure 6A, both MCDW- and DP-derived kw tend to be higher in the young group, and for MCDW-pCASL the difference reached significance (p = 0.0282). CBF is significantly higher in the young group for both sequences (MCDW-pCASL p < 0.0001; DP-pCASL p = 0.0002) (Fig. 6B). In contrast, MCDW-derived Ew doesn’t show significant differences between young and elder subjects, while PSw is increased in the young group (p < 0.0001)(Fig. 6C&D). The reductions in perfusion and BBB parameters with aging can be consistently observed across GM and WM (see Table 2).

**Figure 6.**
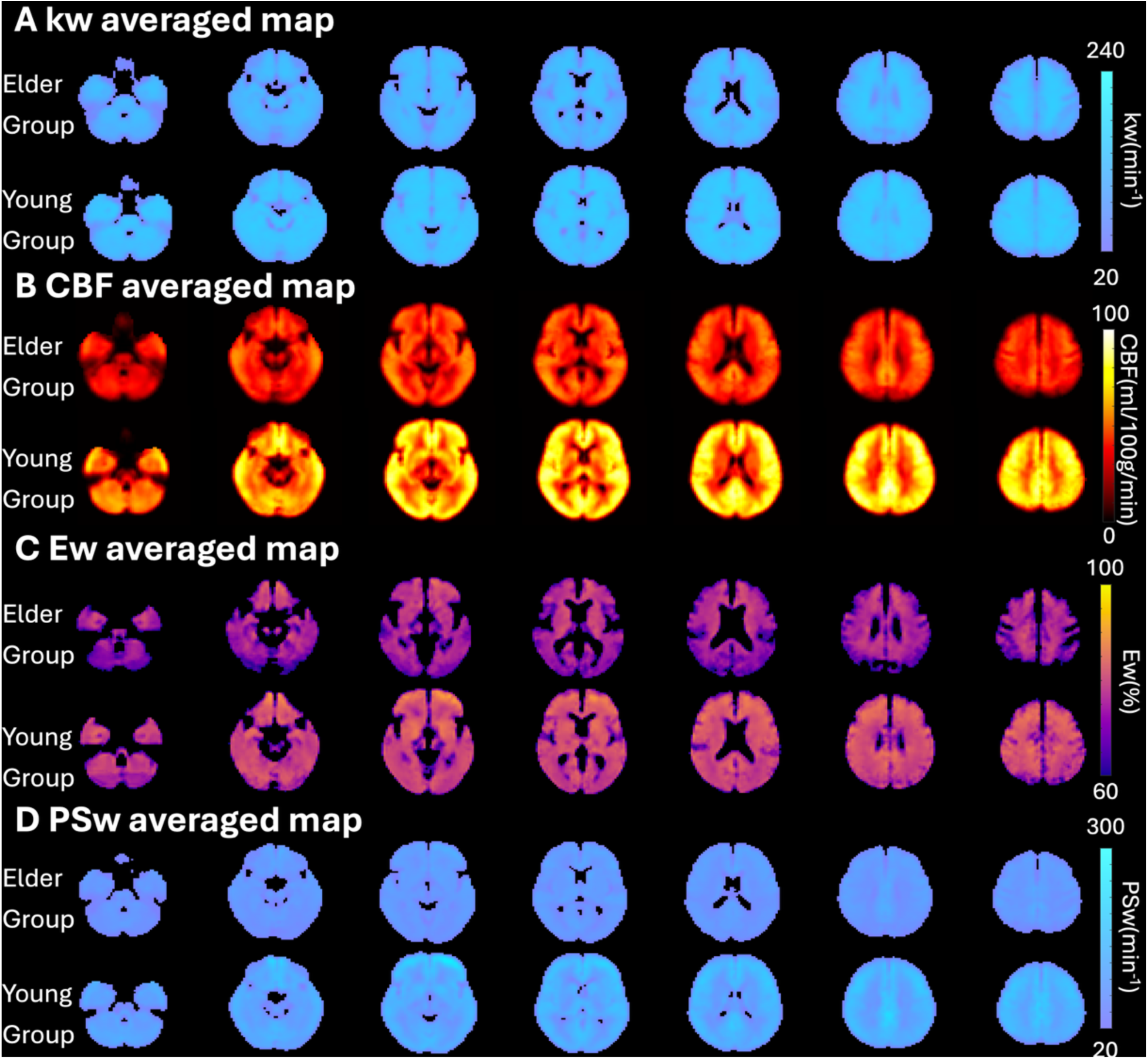
(A) Comparison of kw between young and elderly groups. Blue squares indicate MCDW scans, and red squares represent DP scans. MCDW shows significant differences between the two groups. (B) Comparison of CBF between young and elderly groups. Blue squares indicate MCDW scans, and red squares represent DP scans. Both sequences show significant differences between the groups. (C) Comparison of Ew and PSw from MCDW scans. Green squares represent the elder group, and pink squares represent the young group.

Figure 7 displays group-averaged maps for kw (A), CBF (B), Ew(C), and PSw(D) normalized to the MNI template. The young group exhibits slightly higher kw and notably higher CBF compared with the elder group, consistent with the quantitative results shown in Figure 6.

**Figure 7.**
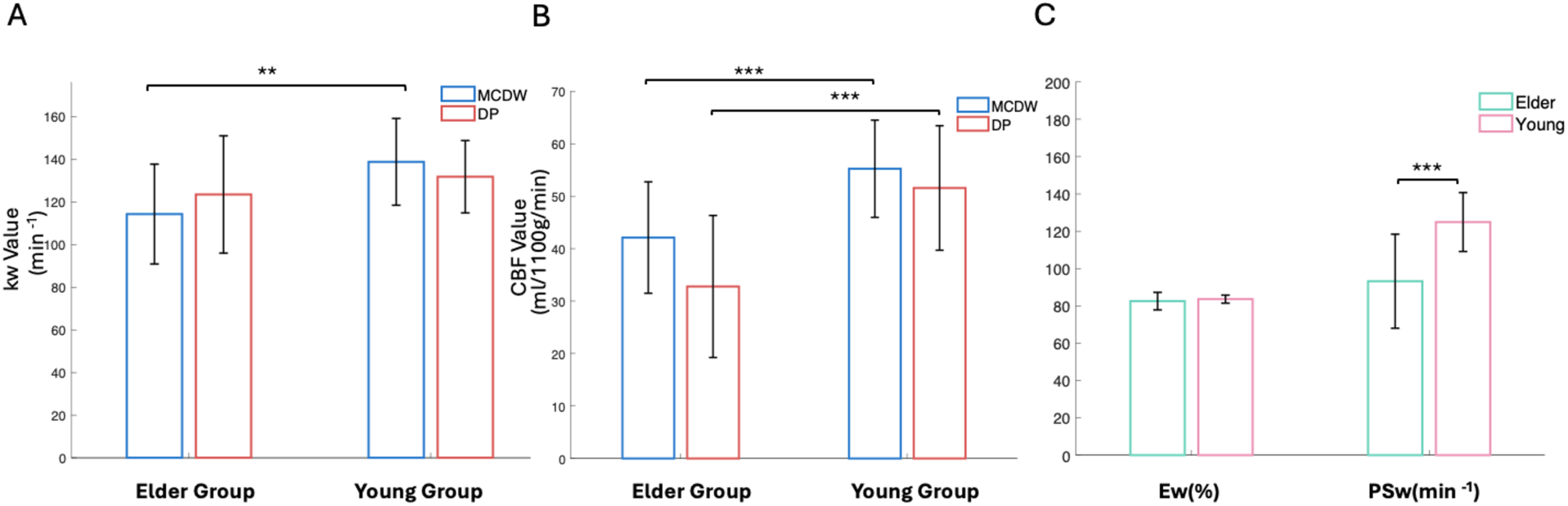
(A–D) Seven representative slices of averaged whole-brain maps from the MCDW-pCASL sequence for the elder (top) and young (bottom) groups. (A) kw maps, (B) CBF maps, (C) Ew maps, and (D) PSw maps.

## 4. DISCUSSION

In this study, we present an accelerated MCDW-pCASL technique that incorporates a low-rank subspace method that enables robust perfusion and BBB water exchange measurements within a clinically feasible scan time of ∼12min. We developed SLIWER to improve the SNR of ASL data, which is particularly beneficial for multi-delay acquisitions and facilitates LRL based strategy for estimating kw from multi-PLD data. In addition, the framework allows estimation of permeability-related metrics (Ew and PSw) from late-PLD signals, providing additional physiological metrics of BBB function beyond kw. This model was not only validated in Monte Carlo simulations, but also by in vivo test–retest experiments, which demonstrated good reproducibility of MCDW-derived kw (ICC = 0.73) and CBF (ICC = 0.75). The permeability metric of PSw also showed relatively stable measurements (ICC = 0.60) despite the low SNR at longer PLDs. Moreover, MCDW exhibited test–retest reliability comparable to that of DP-pCASL. Consistent with previous studies (24), both kw and CBF were lower in the older cohort, supporting the sensitivity of the proposed approach to detect age-related vascular changes.

### 4.1 Low-Rank MRI Denoising

The present study demonstrates that the incorporation of SLIWER denoising with accelerated MCDW-pCASL protocol enables robust quantification of BBB water exchange parameters, while significantly reducing acquisition time from approximately 35 minutes to 12 minutes (21). The kw measurements using the proposed SLIWER framework were highly consistent with those reported in prior MCDW-pCASL studies (previous MCDW: 137.8 ± 12.9 min^-1^; accelerated MCDW: 137.76 ± 20.09 min^-1^), and also demonstrated good to excellent test–retest reliability between repeated scans two weeks apart. These results are consistent with a growing body of low-rank MRI denoising studies, which share the central principle of exploiting redundancy in multidimensional MRI data to separate dominant signal components from noise-dominated components. In diffusion MRI, MP-PCA and NORDIC use PCA/SVD-based locally low-rank modeling to suppress thermal noise while saving diffusion-dependent signal information (31, 32). More recent diffusion MRI studies have further extended this idea by exploiting local patch similarity, diffusion-encoding correlations, and tensor-based multidimensional redundancy for low-rank denoising, including structured low-rank patch-matrix approximation, tensor MP-PCA, and spatial-angular locally low-rank reconstruction for multi-shot DWI (33–35). Similar low-rank principles have also been applied directly to ASL MRI, where multi-channel collaborative low-rank regularization and joint complex low-rank modeling exploit coil/channel, label/control, echo-time, and complex-valued signal correlations for ASL denoising (36, 37).

SLIWER follows this same low-rank denoising principle: it assumes that physiologically meaningful MCDW-pCASL signal evolutions can be represented by a limited number of dominant components, whereas noise contributes less structured components outside the retained subspace. However, instead of relying primarily on local image patches, diffusion-direction redundancy, or coil/channel correlations, SLIWER constructs an SVD-based temporal subspace from combined in-vivo label/control repetitions and Monte Carlo simulated MCDW-pCASL signals. The simulated signals cover the expected range of MCDW-pCASL signal changes across PLDs and diffusion weightings, while the in-vivo data help the basis reflect the actual acquired data on each repetition. This allows SLIWER to suppress less structured noise components while retaining the dominant signal patterns expected from PLD- and diffusion-dependent MCDW-pCASL acquisitions. Taken together, these results, in conjunction with other low-rank-based MRI methods, provide strong support that structured spatiotemporal correlations can be used to overcome the intrinsically low SNR of diffusion-weighted ASL without compromising the accuracy and/or reliability of BBB measurements.

### 4.2 Estimation of Ew and PSw

While the BBB permeability-related metrics in this study showed good test-retest repeatability (PSw ICC = 0.60), the young group averaged values (Ew 83.64 ± 2.28%, and PSw 124.22 ± 19.62 mL/100 g/min) were lower than those reported in previous research. For instance, the WEPCAST study reported Ew of 95.5 ± 1.1% and PSw of 188.9 ± 13.4 mL/100 g/min in healthy participants (38). This difference in signal source may partly be explained since WEPCAST estimates Ew from venous outflow signals, which include both first-pass non-extracted labeled water and delayed tissue-to-vascular re-exchanged labeled water, and derives PSw using the Renkin–Crone relationship, whereas the present MCDW-pCASL method estimates Ew and PSw based on diffusion-weighted ASL signals at the two longest PLDs of 2.4 and 2.7s. These PLDs were selected as a practical compromise among maintaining sufficient SNR, preserving signal reliability, and allowing enough time for water extraction during capillary transit, following prior work showing that tissue fractions measured at comparable late PLDs differed from the model-derived Ew by only approximately 1–3%. However, as these PLDs may not fully meet the timing assumption in all voxels, this limitation may partly contribute to differences in Ew and PSw compared with WEPCAST-derived estimates, which will be discussed in a later section.

### 4.3 Age-Related BBB Water Exchange Changes

In this research, older subjects showed lower whole-brain kw (116.53±19.10 min⁻¹), as well as reduced PSw compared with younger subjects, consistent with prior DP-pCASL findings and studies linking aging and neurodegeneration to impaired BBB water transport (15). Similar age-related declines were observed for CBF, in agreement with values reported using DP-ASL (15). However, the PSw decreasing trend differs from ME-ASL studies, where age-related increases in BBB water permeability have been reported in Alzheimer’s disease rat models and 9.4 T mouse studies, with elevated permeability associated with a 2.1-fold upregulation of aquaporin-4 (AQP4) mRNA expression (39). Interestingly, the directions of kw changes with aging have been shown to be the opposite between ME-ASL and DW ASL (40). These differences may reflect inconsistency in the measured physiological quantity, species, disease status, acquisition design, model assumptions, and physiological compartment being probed, as ASL-based water metrics may reflect perivascular clearance, vascular-to-interstitial exchange, or permeability-related water transport. Nevertheless, water-based BBB metrics—including kw, Ew, and PSw—have been shown to detect pathological alterations earlier than conventional gadolinium-based contrast-agent permeability measures, supporting their potential use as sensitive endogenous biomarkers for early BBB dysfunction (41).

### 4.4 Comparison with DP-pCASL

Our study demonstrated an overall good consistency between MCDW and DP-pCASL techniques. Across subjects, whole-brain CBF values measured by MCDW-pCASL were comparable to those obtained with DP-pCASL, while whole-brain kw estimates remained within a similar physiological range (Table 2). Correlation analysis of the accelerated MCDW-pCASL protocol demonstrated excellent agreement for CBF (ICC of 0.89), but a fair agreement for kw with an ICC of 0.56. These values are comparable to those reported previously for conventional MCDW-pCASL and DP-pCASL, although the lower kw ICC may reflect increased variability associated with the inclusion of older participants.

### 4.5 Limitations

There are several limitations of this study that should be noted. First, the current low-rank subspace database was constructed from a relatively small number of subjects (n = 38), with the majority being older adults (>60 years, n = 32). This imbalance in age distribution may bias the estimation of the kw, given that aging is associated with prolonged transit delays and increased physiological variability. Studies with larger sample sizes and a more balanced age distribution are therefore needed to further validate the robustness of the proposed method. Second, although a five-PLD MCDW-pCASL protocol was used, the longest PLD exhibited a relatively low temporal signal-to-noise ratio (tSNR), which limited its contribution to reliable LRL-based kw estimation. Future protocol optimization should focus on improving the tSNR of late-PLD measurements, and ultra-high-field 7T MRI may provide an opportunity to test whether longer PLDs can improve the robustness of kw, Ew, and PSw estimation (42). Third, Ew and PSw estimation depends on assumptions regarding the capillary transit time τ_c_ and the effective exchange window sampled by the available PLDs. The realistic τ_c_ in each subject/ brain region may vary; and the current PLD settings may not be sufficiently long to fully capture the exchange phase, resulting in biases in calculated Ew and PSw. Future investigation with optimized PLD sampling or independent validation of transit-time assumptions may help reduce uncertainty in Ew and PSw estimation.

## 5. CONCLUSION

MCDW-pCASL combined with the SLIWER framework demonstrates good reliability and accuracy, while providing high spatial resolution and enabling simultaneous multiparametric mapping of cerebral perfusion and BBB water-exchange measures. The method shows overall good agreement with conventional DP-pCASL while achieving shorter scan times and improved image quality. These results highlight the potential of MCDW-pCASL for assessing perfusion and BBB function in clinical populations.

## Data Availability

NA

## ACKNOWLEDGEMENT

This work was supported by the National Institute of Health (NIH) grant R01-NS134712, U01-NS100614, S10-OD032285, R01-EB032169, and RF1AG084072.

## Data and code availability

Data and code can be requested by contacting Dr. Danny Wang at

## Notes

### Competing Interest Statement

The authors have declared no competing interest.

### Author Declarations

All subjects provided written informed consent according to a protocol approved by the Institutional Review Board of the University of Southern California.

